# Associations of conservatism/jumping to conclusions biases with aberrant salience and default mode network

**DOI:** 10.1101/2022.09.29.22280497

**Authors:** Jun Miyata, Akihiko Sasamoto, Takahiro Ezaki, Masanori Isobe, Takanori Kochiyama, Naoki Masuda, Yasuo Mori, Yuki Sakai, Nobukatsu Sawamoto, Shisei Tei, Shiho Ubukata, Toshihiko Aso, Toshiya Murai, Hidehiko Takahashi

**Affiliations:** Department of Psychiatry, Graduate School of Medicine, Kyoto University, Kyoto, 606-8507, Japan; PRESTO, Japan Science and Technology Agency, 4-1-8 Honcho, Kawaguchi, Saitama, Japan; Research Center for Advanced Science and Technology, The University of Tokyo, 4-6-1 Komaba, Meguro-ku, Tokyo, Japan; Brain Activity Imaging Center, ATR-Promotions, 2-2-2 Hikaridai Seika-cho, Sorakugun, Kyoto 619-0288, Japan; Department of Mathematics, State University of New York at Buffalo, NY 14260- 2900, USA; Computational and Data-Enabled Science and Engineering Program, State University of New York at Buffalo, Buffalo, NY 14260-5030, USA; ATR Brain Information Communication Research Laboratory Group, 2-2-2 Hikaridai, Seika-cho, Sorakugun, Kyoto 6190288, Japan; Department of Human Health Sciences, Graduate School of Medicine, Kyoto University, Kyoto 606-8507, Japan; School of Human and Social Sciences, Tokyo International University, Japan; Medical Innovation Center, Kyoto University Graduate School of Medicine, 54 Shogoin Kawahara-cho, Kyoto 6068507 Japan; Laboratory for Brain Connectomics Imaging, RIKEN Center for Biosystems Dynamics Research, Kobe, Japan; Department of Psychiatry and Behavioral Sciences, Graduate School of Medical and Dental Sciences, Tokyo Medical and Dental University, 1-5-45 Yushima, Bunkyo-ku, Tokyo 113-8510, Japan

**Keywords:** default mode network, delusion, energy landscape analysis, meta-independent component analysis, salience, structural covariance analysis

## Abstract

**Background and hypothesis:** While the conservatism bias refers to the human need for more evidence for decision-making than rational thinking expects, people with schizophrenia and delusion require less evidence, known as the jumping to conclusions (JTC) bias. The midbrain-striatal aberrant salience of schizophrenia is postulated to form delusion; however, the association between conservatism/JTC and aberrant salience and their neural correlates are unclear.

**Study design:** Thirty-seven patients with schizophrenia and 33 healthy controls performed the beads task, with large/small numbers of bead draws to decision (DTD) used as an index of conservative/hasty (JTC) decisions, respectively. We performed meta-independent component analysis and thresholded dual regression of resting functional magnetic resonance imaging (MRI) data, and structural covariance network analysis (SCNA) of structural and diffusion MRI data. We investigated interactions between diagnosis and DTD, and main effects of DTD for each imaging modality.

**Study results:** We found no significant interactions. We identified main effects of DTD (positive correlation) for: 1) functional connectivity between the striatum and default mode network (DMN), 2) DMN-like structural SCNA network, and 3) the diffusion SCNA network connecting networks of 1) and 2) (all p < 0.05, family-wise error [FWE] correction). The functional connectivity between the striatum and DMN was also negatively correlated with delusion severity in patients (p < 0.05, FWE), indicating that the greater the anti-correlation, the stronger the JTC and delusion.

**Conclusions:** Our results support a novel concept that conservatism/JTC biases are associated with aberrant salience of schizophrenia and the default brain mode.

## Introduction

Humans form beliefs in response to stimuli in everyday life. This belief formation can be influenced by various cognitive biases. People make conservative inferences for numbers such as likelihood, probability, and frequency, known as the conservatism bias.^1–5^ In a task called the beads task,^6^ beads are repeatedly drawn from one of the two jars with replacement. Jar A contains e.g., 85/15 yellow/blue beads and jar B 15/85. The participants decide from which jar the beads are drawn, and the number of draws needed for decision (DTD) represents the amount of evidence for decision-making. If the first three beads are yellow, Bayes theorem tells us that it is jar A with 85% probability at the first bead, 97% at the second, and 99.5% at the third. However, healthy people need more DTD for decision.^7,8^

Alteration of cognitive biases may lead to abnormal beliefs such as delusion. In the beads task, people with schizophrenia/delusion need less DTD compared to people without these conditions, a difference known as the jumping to conclusions (JTC) bias.^7,9^ Furthermore, some extreme responders require only one bead (DTD = 1) in the 85:15 proportion above.^7^ Recent meta-analyses have reported fewer DTD and more extreme responders among patients with schizophrenia/psychosis, with small to medium effect sizes.^10–13^ JTC is also weakly associated with delusion severity,^10,11^ stable against dopamine agonists/antagonists,^14–17^ and found in patient’s siblings.^18^ Thus, JTC is considered a “trait representing liability to delusion”.^19^

Previous studies on the neural correlates of the conservatism and JTC biases are inconclusive. Functional magnetic resonance imaging (fMRI) studies using the beads task or its modification on healthy individuals^20–25^ (Supplementary Table 1) and patients with schizophrenia/psychosis^26–28^ (Supplementary Table 2) reported multiple brain regions. This may be because 1) the beads task is not suitable for repeated trials in the scanner,^29^ 2) modification could change the task nature,^30^ and 3) different statistical procedures such as contrasts and statistical thresholds could affect the results. Only one structural MRI^31^ and no diffusion MRI study have been reported.

Elucidation of the neural mechanisms of JTC and delusion requires an integrated understanding of multiple brain regions/networks. Independent component analysis (ICA) can identify intrinsic functional networks using resting fMRI in a data-driven manner and “structural covariance networks” using structural^32,33^ and diffusion MRI.^34,35^ The dopaminergic aberrant salience-related networks^36^: the medial temporal lobe network (MTLN, the bilateral hippocampus) and basal ganglia network (BGN, the midbrain and striatum) are important group of networks. Glutamatergic hyperactivity in the hippocampus causes a hyper-dopaminergic state in the midbrain and striatum,^37^ attributing excessive salience to ordinary stimuli and leading to delusion.^38^

Other important networks, including “triple networks”, i.e., the salience network (SN, the anterior cingulate cortex [ACC] and insula), frontoparietal network (FPN, the bilateral frontal and parietal cortices), and default mode network (DMN, the anterior and posterior medial cortices and lateral parietal cortices).^39–41^ have been associated with various psychiatric disorders including schizophrenia.^41^

This study investigated the functional and structural neural correlates of conservatism and JTC biases using conventional “one-shot” beads task outside the scanner and ICA of resting functional, structural, and diffusion MRI. To investigate multiple aspects of functional connectivity, we performed within- and between-network connectivity analyses and dynamic (time-varying) functional network analysis. We hypothesized that conservatism/JTC biases were associated with aberrant salience-related networks.

## Methods

### Participants

Thirty-seven patients with schizophrenia (SCZ) and 33 age- and sex-matched healthy controls (HC) were recruited at Kyoto University Hospital. Patients’ general psychopathology was assessed using the Positive and Negative Syndrome Scale (PANSS).^42^ The severity of delusional belief, ranging from subclinical delusional ideation in HC and delusions in SCZ, was assessed using the Peters et al. delusions inventory (PDI).^43^ The premorbid IQ was measured by the Japanese version of the National Adult Reading Test short form (JART-25)^44,45^. All patients were administered antipsychotics (Supplementary Methods).

After receiving a complete description of the study, all participants provided written informed consent. The study design was approved by Kyoto University Graduate School and Faculty of Medicine, Ethics Committee.

### Beads task

The participants were presented with jars A and B, containing 80/20 blue [b]/yellow [y] and 20/80 b/y beads, respectively. Beads were drawn from one of the jars repeatedly one by one, with a maximum draw of 20, following the order of yyybyyyyybbyyyyyyyyb (Figure 1). The participants were asked at each draw to 1) decide from which jar the beads were drawn, or 2) request another draw. DTD was recorded as the index of conservatism/JTC tendency. Extreme responders were defined as participants with DTD = 1^46^ (the probabilities of jar A were 80%, 94.1%, and 98.5% at the first, second, and third draws, respectively).

**Figure 1.**
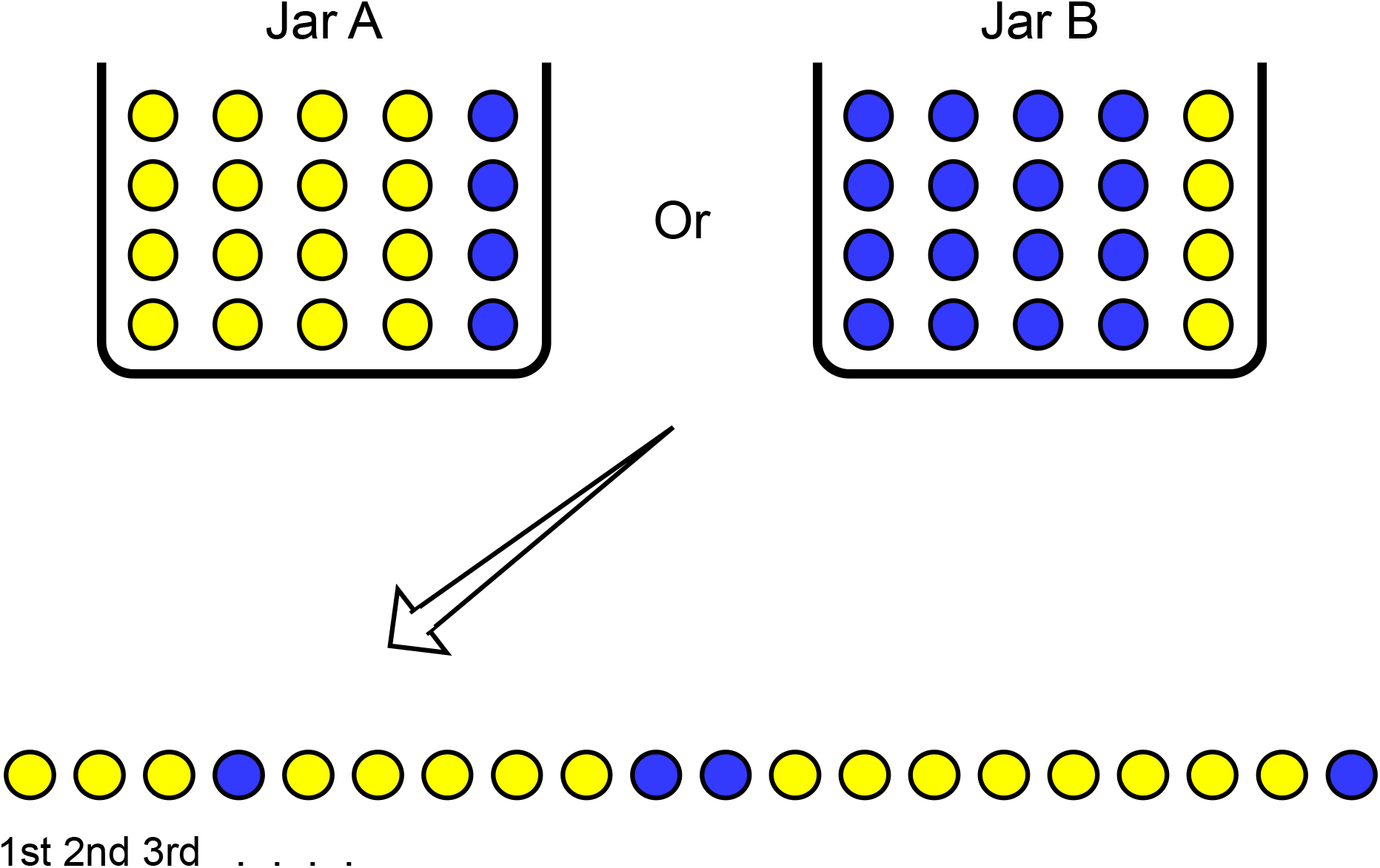
Beads task. Beads were drawn from jar A (80 yellow and 20 blue beads) or B (20 blue and 80 yellow beads), repeatedly one by one, with a maximum draw of 20, following the presented order. The participants were asked at each draw to 1) decide from which jar the beads were drawn, or 2) request another draw. The number of draws to decision (DTD) was recorded as the index of conservatism/JTC tendency.

### MRI acquisition

The resting fMRI and structural and diffusion MRI data were acquired on a 3T scanner (Siemens, Erlangen, Germany). For resting fMRI, participants with excessive head motion (mean framewise displacement^47^ >0.5 mm) were excluded. The details including the sequences and parameters are described in Supplementary Methods and Supplementary Table 3.

### Resting fMRI analyses

#### 1) Preprocessing

The details of resting fMRI preprocessing are described in Supplementary Methods. In addition to the standard preprocessing steps, we performed ICA-based denoising^48^ of each participant’s resting fMRI data and removed non-neuronal signals (head motion, physiological noise such as pulsation and respiration, susceptibility-induced noise, and machine-derived noise).

As an index of data quality, we calculated the temporal signal-to-noise ratio (tSNR) at each voxel as the mean/SD of the time course averaged across the whole brain using QAscript.^49^

#### 2) Meta-ICA using a large dataset

We performed meta-ICA,^50,51^ which has higher robustness and reproducibility for component estimation than conventional single-shot group ICA (gICA).^52^ The procedures are described in our previous study (doi:10.1101/2021.10.02.21264326). Briefly, 40 patients and 40 controls were randomly selected from a large dataset of 209 patients with psychosis and 279 unaffected individuals, which included 34 participants with SCZ and 28 HC in this study. Temporal concatenation gICA was then performed using the MELODIC ^53^ (version 3.15) toolbox of FSL (version 6.0.3) (http://www.fmrib.ox.ac.uk/fsl). This step was repeated 25 times, and a gICA was performed on the concatenation of the 25 gICA results.^52^ We repeated these steps, incrementing the total independent component (IC) number from 20 by five. When the total IC number was 80, we identified the following 11 networks of interest (NOIs): the MTLN, midbrain-thalamic and striatal parts of the basal-ganglia network (BGN-MbThal and BGN-Str), ACC and insular parts of the SN (SN-ACC and SN-Ins), right and left FPN, and four subnetworks of DMN (DMN1–4) (Figure 2a).

**Figure 2.**
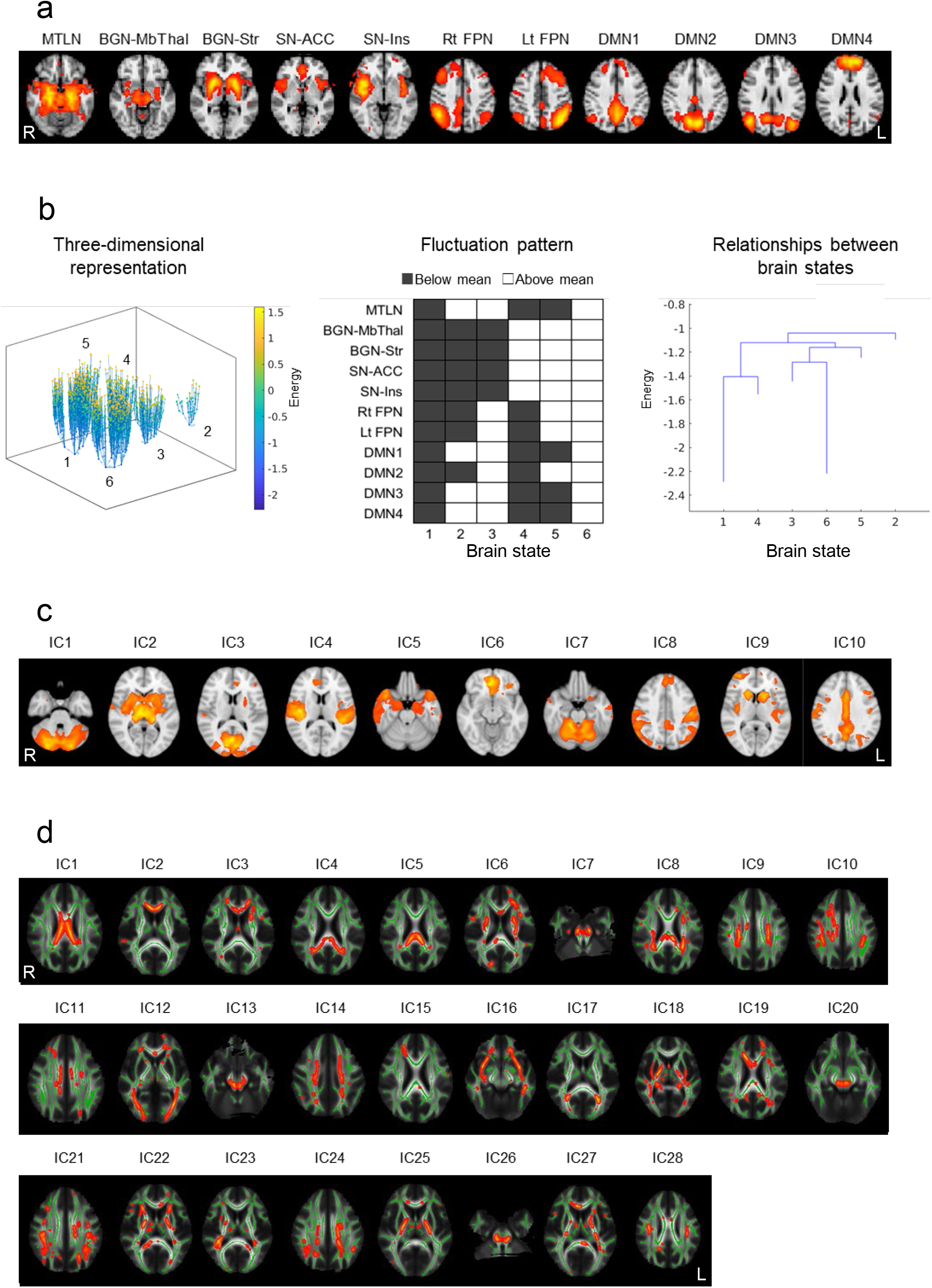
Group-level network findings. a. Eleven NOIs of resting fMRI created by meta-ICA. b. Time courses of 11 NOIs aggregated into six brain states (BS) (left). The fluctuation pattern of local minima of each BS is shown in the middle. BS1 and BS6 were more stable than BS2, BS3, BS4, and BS5 (right). c. NOIs of structural MRI (gray matter) created by SCNA. d. NOIs of diffusion MRI (white matter) created by SCNA. **Abbreviations** ACC, anterior cingulate cortex part; BGN, basal ganglia network; DMN, default mode network; fMRI, functional magnetic resonance imaging; FPN, frontoparietal network; ICA, independent component analysis; Ins, insula part; MbThal, midbrain-thalamus part; Lt, left; MTLN, medial temporal lobe network; NOI, network of interest; Rt, right; SCNA, structural covariance network analysis; SN, salience network; Str, striatum part.

### 2 Thresholded dual regression

Next, using the ICs (spatial maps) created in meta-ICA, we performed thresholded dual regression on our dataset.^54^ First, we regressed each participant’s preprocessed data against the spatial maps of meta-ICA to obtain participant-specific time courses for each IC. Second, we regressed the preprocessed data against the participant-specific time courses to obtain participant-specific spatial maps for each IC. Third, we applied mixture model thresholding to participant-specific spatial maps.^53^ Fourth, we regressed the preprocessed data against these thresholded spatial maps to obtain participant-specific time courses. The time courses at the fourth and first steps represented the regions inside the networks and the whole brain (including the regions outside the networks), respectively.^54^

We used the spatial maps from the second step for statistical analysis of within-network connectivity. For between-network connectivity, partial correlations between the time courses at the fourth step were calculated using FSLNets 0.6.3 (https://fsl.fmrib.ox.ac.uk/fsl/fslwiki/) to control for the other nine NOIs. Among the 55 NOI-pairs, we selected 12 that passed the pre-masking at |t| ≥ 8. Correlation coefficients were Fisher-transformed into Z-scores and used for statistical analysis.

### 3) Energy landscape analysis

Next, to analyze brain dynamics, we performed energy landscape analysis (ELA) using ELA toolbox 2.0 (https://github.com/tkEzaki/energy-landscape-analysis).^55–60^ Briefly, time course fluctuations at each time point were binarized as 1 and -1, depending on whether the value of a time point was above or below the mean of each time course. The 11 NOIs corresponded to 2048 (2^11^) possible fluctuation patterns at each time point. We concatenated the binarized time-series data of all participants (175 × 70 = 12250 total time points) and calculated the frequency (probability) of each fluctuation pattern. We then fitted the pairwise maximum entropy model to the data to calculate the “energy” of each activity pattern. Figure 2b (left) shows a three-dimensional view of the “energy landscape,” in which each plot represents each fluctuation pattern. This energy landscape consisted of six brain states (BS). The fluctuation patterns of the local minima (frequent, low-energy patterns) are shown in Figure 2b (middle). BS1 and BS6 showed lower energies and, hence, higher frequencies than BS2, BS3, BS4, and BS5 (Figure 2b, right). Thus, we calculated the 1) frequency of (BS1 + BS6), 2) transition rate between B1 and B6, and 3) transition rate between (BS1 + BS6) and (BS2 + BS3 + BS4 + BS5) for the statistical analysis.

### Structural MRI analysis

We used CAT12 (http://www.neuro.uni-jena.de/cat), an extension of SPM12 (https://www.fil.ion.ucl.ac.uk/spm/software/spm12), to segment the structural data into gray matter, white matter, and cerebrospinal fluid, and performed normalization. The gray matter segment was then resampled into 1.5 mm, Jacobian-modulated for nonlinear effects only, and smoothed by 8-mm full-width at half-maximum. The voxel value of this gray matter “density” map reflected local gray matter volume. We did not apply Jacobian modulation for linear effect since it was not suitable for the following analysis.

Next, we performed structural covariance network analysis (SCNA).^32^ ICA by MELODIC/FSL was performed on the preprocessed gray matter segment from 33 SCZ and 33 HC participants, treating them as four-dimensional data. The number of ICs was set at 10 according to previous studies.^61,62^ Since there is less consensus on the classification/characteristics of gray matter covariance networks compared to those of resting fMRI networks, we used all 10 ICs as the gray matter NOIs (Figure 2c). We then regressed the four-dimensional participant data against the 10 ICs to obtain the loading values for each NOI in each participant, as an index of gray matter covariance strength, which was used for the statistical analysis.

### Diffusion MRI analysis

We performed noise level estimation and denoising on raw diffusion MRI data based on random matrix theory, using dwidenoise in MRtrix3 (https://www.mrtrix.org/). The data were then corrected for eddy current, head motion, and susceptibility-induced distortion using the dti_preprocess tool (http://www.bic.mni.mcgill.ca/~thayashi/dti.html). Then fractional anisotropy (FA), an index of white matter integrity, was calculated by DTIFIT/FSL, and FA maps were “skeletonized” using the tract-based spatial statistics (TBSS) algorithm.^63^ The skeletonized FA maps of all participants were entered into the SCNA as in the structural MRI analysis. The number of ICs was set to 28 based on previous studies,^64,65^ and all ICs were used as white matter NOIs (Figure 2d). Each participant’s loading values were used for the statistical analysis as an index of white matter covariance strength.

For data quality assurance, we calculated mean tSNRs using nine b=0 data.^49^

## Statistical analysis

Demographic and clinical data were analyzed using SPSS 27 (IBM). Permutation-based non-parametric inference was performed for imaging analyses in PALM^66^ (version 116) of FSL. Using the general linear model (GLM) we first tested the interaction between diagnosis and DTD. If no interaction was found, we then tested the main effect of DTD (positive or negative correlation) on the resting fMRI, structural, and diffusion MRI indices. Age, sex, mean tSNR (for resting fMRI and diffusion MRI), and premorbid IQ was used as covariates since IQ could affect DTD.^67,68^ The statistical threshold was p<0.05, correcting for:

A) voxels (within-network analysis only): threshold-free cluster enhancement (TFCE)^69^ within each NOI,

B) contrast number: two contrasts (positive and negative interactions/main effects), and

C) network/index numbers: 11 NOIs for within-network analysis, 12 NOI-pairs for between-network analysis, three indices for ELA (one frequency and two transition rates), and 10 gray matter and 28 white matter NOIs.

If we found significant interactions or main effects, we tested the correlations between imaging indices and delusion severity (PDI total score) for patients with SCZ, with age, sex, premorbid IQ, and tSNR as covariates. The statistical threshold was p<0.05, corrected for contrasts, as well as for networks/indices if more than two networks/indices were found to be significant.

We similarly checked the effect of medication (chlorpromazine equivalent (CP eq)).

### Relationships between fMRI, gray matter, and white matter findings

We investigated the anatomical and correlational relationships between significant fMRI, gray matter, and white matter findings. We investigated the correlation between all pairs of imaging indices using the same covariates as above. The statistical threshold was p<0.05, corrected for contrasts.

## Results

### Demographic and clinical data

Table 1 demonstrates the trend of lower premorbid IQ in SCZ compared to HC. The PDI total score was significantly higher in SCZ. The DTD did not differ between groups. However, SCZ showed a trend of higher number of extreme responders. The tSNR for fMRI and diffusion MRI did not differ between groups.

**Table 1.** Demographic and clinical data.

|  | SCZ (n=37) | HC (n=33) | Statistics | DoF | P value |
| --- | --- | --- | --- | --- | --- |
|  | Mean (SD) |  |  |  |  |
| Age (years) | 37.8 (9.4) | 34.4 (7.6) | $t = -1.70$ | 67.33 | 0.09 |
| Age range (years) | 24–56 | 21–57 |  |  |  |
| Sex (female/male) | 20/17 | 13/20 | $\text{Chi}^2 = 1.50$ | 1 | 0.22 |
| Premorbid IQ (JART) | 104.1 (10.5) | 108.6 (8.0) | $t = 1.98$ | 68 | 0.052 |
| Smoker/non-smoker | 4/33 | 3/28 | $\text{Chi}^2 = 0.023$ | 1 | 0.88 |
| Duration of illness | 12.7 (8.1) |  |  |  |  |
| Medication (CPeq mg) | 556.6 (477.1) |  |  |  |  |
| PANSS total | 57.9 (18.2) |  |  |  |  |
| positive | 14.1 (5.3) |  |  |  |  |
| negative | 15.1 (5.7) |  |  |  |  |
| general | 28.7 (10.2) |  |  |  |  |
| PDI total | 106.9 (67.8) | 21.2 (22.1) | $t = -7.17$ | 42.97 | < 0.001 |
| DTD | 4.6 (2.4) | 4.5 (2.1) | $t = -0.26$ | 68 | 0.80 |
| Extreme responder | 4 | 0 | $\text{Chi}^2 = 3.78$ | 1 | 0.052 |
| tSNR (fMRI) | 1530.4 (400.2) | 1420.5 (341.6) | $t = -1.23$ | 68 | 0.22 |
| tSNR (diffusion MRI) | 28.2 (3.9) | 28.9 (5.3) | $t = 0.67$ | 68 | 0.51 |

The DTD was not significantly significantly correlated with the PDI total score (SCZ: r = 0.003 and p = 0.97; HC: r = 0.26 and p = 0.15) or medication (r=-0.16 and p=0.33).

### Resting fMRI and structural and diffusion MRI analyses

Within-network functional connectivity did not show significant interactions between diagnosis and DTD or main effects of DTD (p>0.05, corrected for voxels by TFCE, two contrasts, and 11 networks).

Between-network functional connectivity did not show a significant interaction but showed a significant main effect of DTD (positive correlation) for connectivity between the BGN-Str and DMN2 (the precuneus) (p=0.02, corrected for two contrasts and 12 NOI-pairs. Figure 3a).

**Figure 3.**
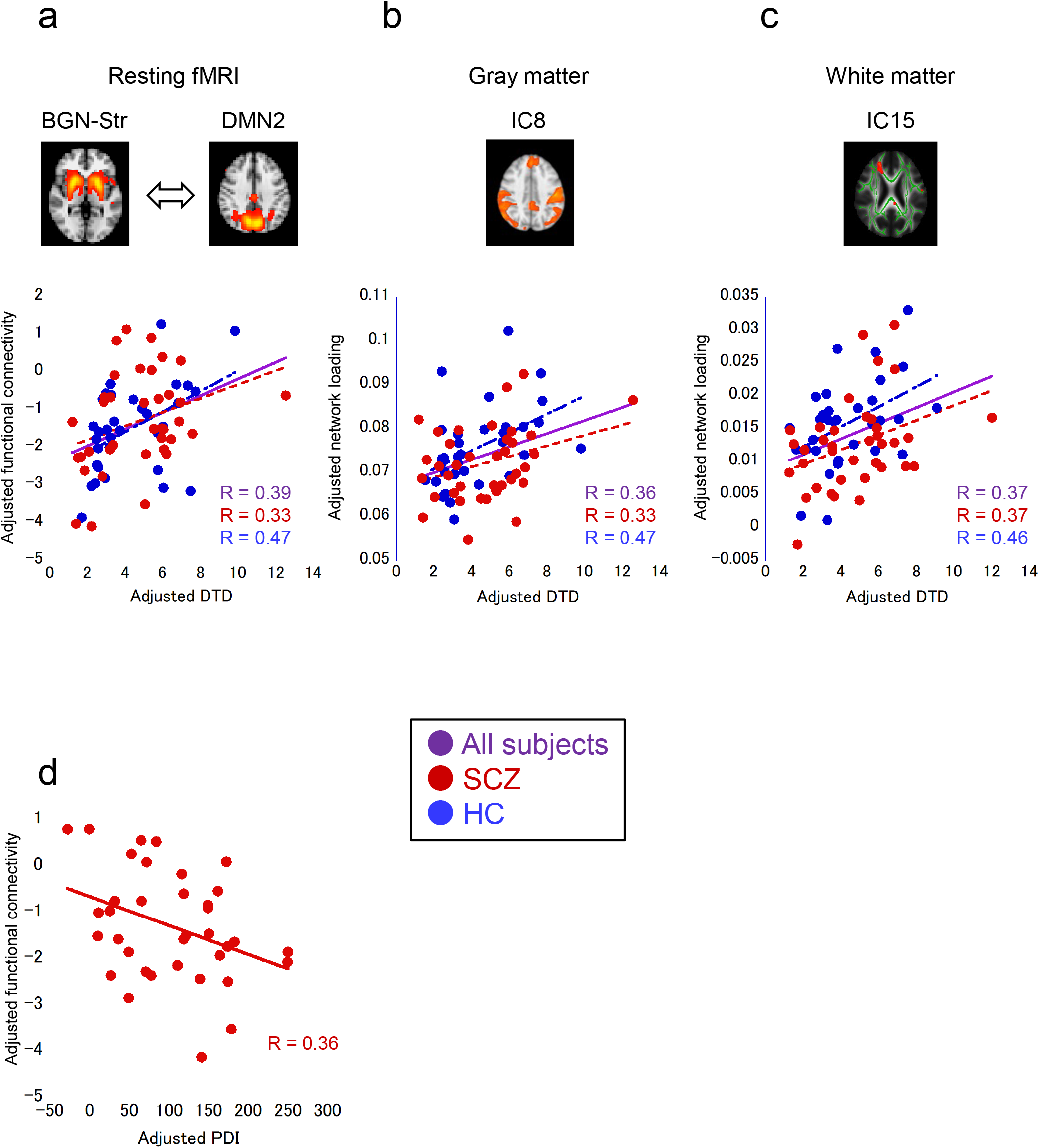
Associations between DTD, connectivity, and delusion. DTD and functional connectivity values were adjusted for age, sex, JART, and tSNR (fMRI and white matter). a. Significant main effect of DTD on the functional connectivity between the BGN-Str and DMN2. p<0.05, corrected for two contrasts and 12 network-pairs. b. Significant main effect of DTD on the network loading value for the DMN-like gray matter covariance network (IC8). p<0.05, corrected for two contrasts and 10 networks. c. Significant main effect of DTD on the network loading value for a white matter covariance network (IC15). p<0.05, corrected for two contrasts and 28 networks. d. Significant correlation between delusion severity measured by PDI and functional connectivity between the BGN-Str and DMN2 in SCZ. p<0.05, corrected for two contrasts. **Abbreviations** BGN-Str, striatum part of the basal ganglia network; DMN, default mode network; DTD, draws to decision; fMRI, functional magnetic resonance imaging; HC, healthy controls; IC, independent component; JART, Japanese Adult Reading Test; PDI, Peters et al delusions inventory; SCZ, patients with schizophrenia; tSNR, temporal signal-to-noise ratio.

The ELA indices (i.e., frequency and transition rate of brain states) did not show significant interactions or main effects (p>0.05, corrected for two contrasts and three indices).

The loading values for the gray matter covariance network did not show significant interactions but showed a significant main effect (positive correlation) for a DMN-like covariance network containing the medial prefrontal and posterior cingulate cortices and bilateral dorsolateral parietal cortices (IC8, p=0.008, corrected for two contrasts and 10 NOIs. Figure 3b).

The loading values for the white matter covariance network did not show significant interactions but showed a significant main effect (positive correlation) for a covariance network (IC15 p=0.03, corrected for two contrasts and 28 NOIs. Figure 3c).

### Correlation between imaging indices and delusion severity/antipsychotic

#### medication dose

The functional connectivity between the BGN-Str and DMN2 was negatively correlated with the PDI total score for SCZ (p=0.02, corrected for two contrasts. Figure 3d). The PDI total score and covariance loading values of the gray matter (IC8, p=0.38, corrected for contrasts) and white matter (IC15, p=0.63, corrected for contrasts) showed no correlations.

Medication was not correlated with functional connectivity between the BGN-Str and DMN2 (p=0.32, corrected for contrasts), but showed trend-level negative correlations for the loading values of gray matter (IC8) and white matter (IC15) (p=0.051 and 0.08, respectively, corrected for contrasts, Supplementary Figure 1).

#### Relationships between fMRI, gray matter, and white matter findings

The two resting-fMRI (BGN-Str and DMN2), gray matter (IC8), and white matter (IC15) NOIs were adjacent (Figure 4a). We observed a significant positive correlation between gray matter and white matter loading values (p=0.003, corrected for two contrasts. Figure 4b).

**Figure 4.**
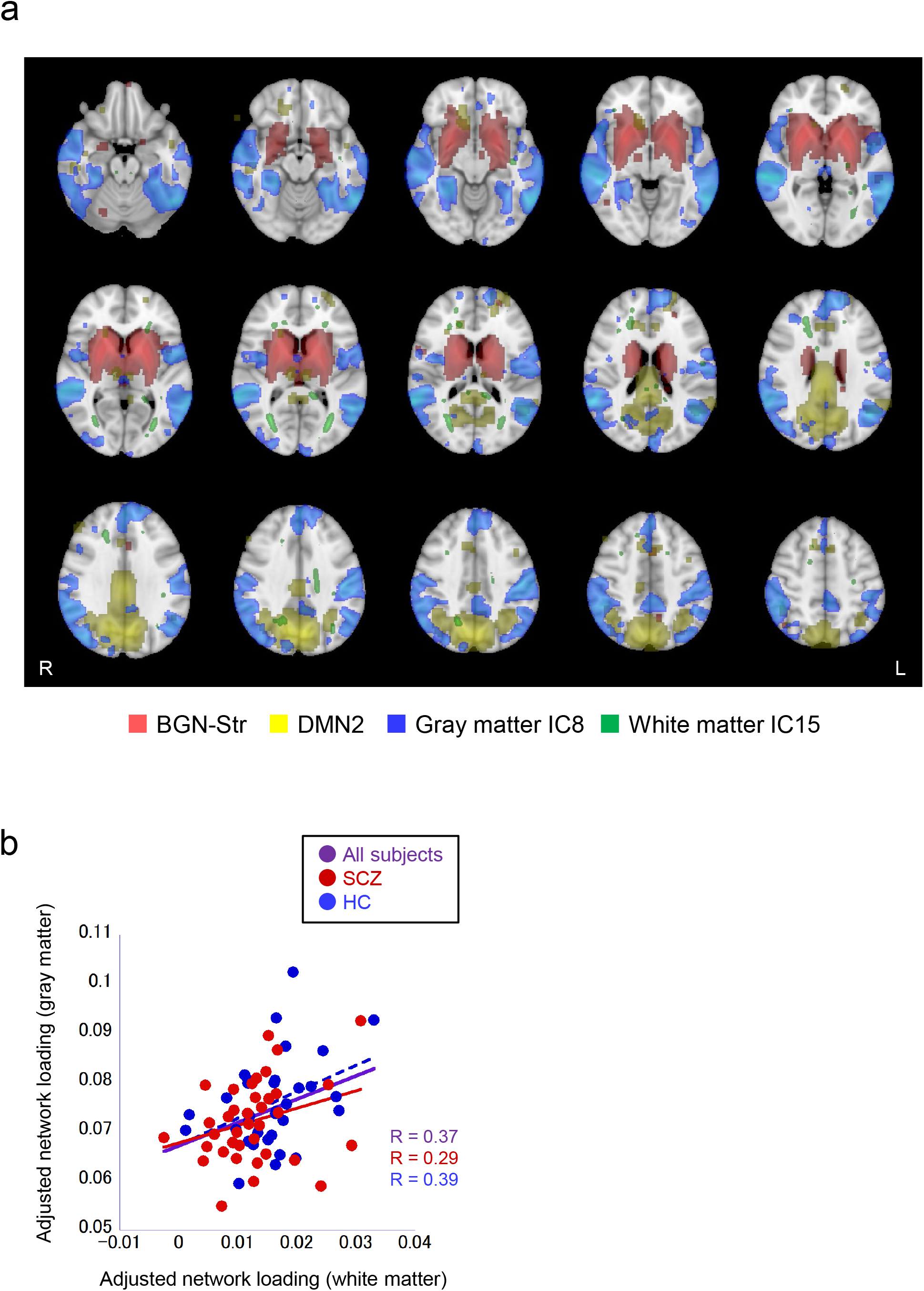
Associations between functional, gray matter, and white matter NOIs. a. The functional NOIs (red: BGN-Str, yellow: DMN2) and gray matter (blue: IC8) and white matter (green: IC15) covariance networks are adjacent. b. Gray matter and white matter loading values were significantly correlated (p<0.05, corrected for two contrasts). The loading values were adjusted for age, sex, JART, and tSNR (white matter). **Abbreviations** BGN-Str, striatum part of the basal ganglia network; DMN, default mode network, HC, healthy controls; JART, Japanese Adult Reading Test; NOI, networks of interest; SCZ, patients with schizophrenia; tSNR, temporal signal-to-noise ratio.

## Discussion

The main study findings were:

1) The functional networks and gray matter/white matter covariance networks around the striatum and DMN were associated with conservatism and JTC biases.

2) The functional connectivity between the striatum and DMN was also associated with delusion severity in SCZ.

3) The functional, gray matter, and white matter networks were anatomically adjacent, with the last two also showing statistical associations.

Collectively, we identified converging evidence of the functional, gray matter, and white matter correlates of the conservatism and JTC biases and delusion, as discussed in detail in the following sections.

### Functional connectivity and associations among conservatism/JTC, delusion, aberrant salience, and the DMN

Most participants showed a negative correlation coefficient between the BGN-Str and DMN2 (the precuneus), indicating the anti-correlation of these two networks. Stronger anti-correlation was associated with stronger JTC, while weaker anti-correlation was associated with stronger conservatism. Previous fMRI studies using beads task reported activation in the striatum^27^ and precuneus/DMN.^21,23,25,26^

The stronger the anti-correlation, the stronger the delusion severity in patients with SCZ, providing the first direct support for the neural link between JTC and delusion. This result also supports the link between the JTC and the aberrant salience hypothesis^38^ featuring the striatum, both of which explain delusion formation, and is consistent with a behavior study with a modified beads task that also indicated this link.^70^

A previous fMRI study using a task like the beads task showed that the precuneus was activated proportionally to the uncertainty level.^71^ Another study revealed that larger DTD (conservatism) was associated with more analytic rather than intuitive cognitive style.^72^ Thus, the precuneus/DMN may link conservatism bias and dual process theory.^73^

The association of DTD with between-network but not within-network connectivity may indicate that conservatism/JTC requires interactions between multiple brain networks. Moreover, the lack of association of DTD with temporary-varying analysis of ELA may be consistent with the concept of JTC as a “trait” (rather than state) representing a liability to delusion. The use of all 11 NOIs to construct the energy landscape may also have blurred the potential association by including irrelevant NOIs.

### Gray matter/white matter covariance and their association with functional connectivity

This is the first study to report the gray matter and white matter structural correlates of conservatism and JTC biases. The gray matter covariance network (IC8) belonged to the DMN, further supporting the findings of the functional connectivity analysis above.

The white matter covariance network (IC15) was mainly located in the frontal/prefrontal regions, as well as anatomically adjacent to the DMN2 and gray matter IC8 (Figure 4a). This anatomical adjacency, as well as the positive correlations between conservatism/JTC and the functional, gray matter, and white matter networks, may indicate shared mechanisms among these modalities. This is consistent with our previous findings of associations between gray matter and white matter alterations in schizophrenia.^74–76^

The trend-level association between gray matter and white matter covariance and medication indicated that patients with lower covariance (“structural connectivity”) tended to have stronger JTC and received higher-dose antipsychotics. This result is consistent with the discussion above.

### Methodological strengths

The methodological strengths of this study include intensive fMRI data denoising using ICA-denoising, robust estimation of functional networks using meta-ICA on a large sample data set, thresholded dual regression to accurately estimate network time course, SCNA to identify structural covariance networks of gray matter and white matter, and full multiple-comparison correction, all of which contributed to the increased reproducibility of this study.

## Limitations

First, our sample size was not large and replication of our findings with larger sample size is necessary. The sample size might also be the reason of no significant group differences in DTD or the number of extreme responders. However, these are not inconsistent with those of a previous meta-analysis, in which 15 of 33 studies reported no significant difference in DTD, and seven of 22 studies for extreme responders.^11^ Second, all patients with SCZ were on antipsychotics; thus, we could not fully investigate the effects of medication. Third, we did not perform susceptibility-induced distortion correction for resting fMRI data since this was not performed in the large dataset used for meta-ICA.

## Conclusions

ICA of multi-modal MRI data revealed functional, gray matter, and white matter neural correlates of conservatism/JTC cognitive biases and delusion, which were associated with aberrant salience and default brain mode. Investigations of cognitive biases using multi-modal approaches will elucidate the pathophysiology of schizophrenia and delusion and the mechanisms of human cognition.

## Data Availability

All data produced in the present study are available upon reasonable request to the authors and after MTA.

## Acknowledgments

We thank the patients and volunteers for their participation. We also thank the lab members for their contributions and COCORO consortium for their cooperation.

This work was supported by grants from KAKENHI (Japan Society for the Promotion of Science and the Ministry of Education, Culture, Sports, Science and Technology) (grant numbers 26461767, 17H04248, 18H05130, 20H05064, 20K21567, 19H03583, and 21K07544); the Japan Agency for Medical Research and Development Brain/MINDS & beyond studies (grant numbers JP18dm0307008 and JP21uk1024002); the Japan Science and Technology Agency (JST) for the financial support (under Grant No. JPMJMS2021);

a Novartis Pharma Research Grant;

the SENSHIN Medical Research Foundation;

the Uehara Memorial Foundation;

the Kyoto University Global Frontier Project for Young Professionals;

and the Takeda Science Foundation.

All authors have declared that there are no conflicts of interest concerning the subject of this study.

