## Supplementary material for "Associations of conservatism/jumping to conclusions biases with aberrant salience and default mode network"

### Supplementary Methods

#### Participant details

All participants were Japanese or native Japanese-speaking east Asians. The diagnoses were made based on the Structured Clinical Interview for Diagnostic and Statistical Manual of Mental Disorders (DSM-IV) Axis I Disorders (SCID). Patients with schizophrenia (SCZ) included those with a diagnosis of schizophrenia, schizoaffective disorder, or schizophreniform disorder. Patients were not comorbid with any other Axis I disorders.

The healthy control (HC) participants had no first-degree relatives with psychotic disorders. The exclusion criteria for all individuals included a history of head trauma, neurological illness, serious medical or surgical illness, and substance abuse.

#### MRI acquisition details

The scanner was equipped with a 40 mT/m gradient and a receiver-only eight-channel phase-array head coil.

During the resting fMRI scanning, the participants were instructed to visually concentrate on a fixation cross in the center of the screen while thinking nothing at all.

All MRI images were visually checked, and participants with gross artifacts or anatomical anomalies were excluded.

#### Resting fMRI preprocessing

Head-motion correction was performed using SPM8 (Wellcome Department of Cognitive Neurology, London, United Kingdom) running on MATLAB (MathWorks), followed by spatial normalization to the MNI template brain using the T1 anatomical image.

ICA-based denoising was performed for each participant to remove non-neuronal signals (head motion, physiological noise such as pulsation and respiration, susceptibility-induced noise, and machine-derived noise) from the fMRI data (for details, please refer to Aso et al, 2017 [1]). The fMRI images were fed into MELODIC/FSL (http://www.fmrib.ox.ac.uk/fsl) with automatic component number estimation. Instead of selecting several participants as a training sample for machine-learning and classifying the components of other participants into neuronal and noise components [2,3], we classified all participants' components based on three objective criteria: 1) high-frequency power ratio (>0.1 Hz up to 0.2 Hz or the Nyquist frequency for the TR) relative to <0.1 Hz, 2) non-gray matter involvement index, and 3) slice dependency index calculated as the ratio of within- and between-slice spatial high-frequency components. Components with high-frequency power ratio (HF), high slice dependency index (SL), or high non-gray matter index (NB) were classified as noise, while components with neuronal origin were preserved.

After ICA-denoising, the resting fMRI data were normalized into the MNI 152 space using the unified segmentation algorithm of SPM8. The data were then re-sampled into 3 mm isotropic voxels and smoothed using a Gaussian kernel of full width at a half maximum of 5 mm. We discarded the first five volumes for signal stabilization.

**Supplementary Table 1. fMRI studies applying the beads task or its modification in healthy participants.**

| Study | Beads task | Response | Reward /loss | Feedback | Contrast | Brain region |
|---|---|---|---|---|---|---|
| Blackwood et al, 2004 | Classical | DTD | No | No | Bead task - control [a] | Lt cerebellum, PMC, IPL, and visual cortex |
| Furl & Averbeck 2011 | Classical | DTD | Yes | Yes | Decision - draw [b] | ACC |
| | | | | | | Bil insula, parietal cortex, and Str |
| | | | | | | Lt GB, Thal, and MB |
| | | | | | Draw - decision [b] | Precuneus and PCC |
| | | | | | | Lt SFG and Lt TPJ |
| | | | | | | Rt visual cortex |
| | | | | | Correlation with DTD [c] | Rt DLPFC and parietal cortex |
| | | | | | Decision - draw (parametric modulation by costed action value) [b] | Rt insula |
| Esslinger et al, 2013 | Modified (Fish task) | DTD | Yes/No | Yes/No | Bead task - control [d] | PCC and ACC |
| | | | | | | Bil cerebellum and Thal |
| | | | | | | Lt IFG, precentral gyrus, MFG/SMA, insula, and lingual gyrus |
| | | | | | | Rt MFG, IFG, and medial frontal gyrus/SMA, fujiform gyrus, SPL, and MB |

| | | | | | | |
|---|---|---|---|---|---|---|
| Whitman et al, 2013 | Modified (Fish task) | Probability estimation | No | No | Confirmatory - disconfirmatory [e] | Component 1: dACC, Bil parietal and lateral occipital cortices |
| | | | | | | Component 2: DMN (deactivation) |
| | | | | | Bead task - control [e] | None |
| Demanuele et al, 2015 | Modified (Fish task) | DTD | No | No | ROIs vs. control areas [f] | ROIs (ACC, Bil DLPFC,and OFC) |
| Andreou et al, 2018a | Modified (Box task) | DTD | No | No | Decision - draw [g] | ACC and precuneus |
| | | | | | | Bil parietal cortex, insula, lingual gyrus, and cuneus |
| | | | | | | Rt DLPFC, PFC, MB, and cerebellum |
| | | | | | Draw - decision [g] | Ventral ACC and PCC |
| | | | | | | Lt angular gyrus |
| | | | | | | Rt insula |
| | | | | | PPI from Lt visual cortex (decision - draw) [h] | Ventral ACC |
| | | | | | | Bil visual cortex |
| | | | | | | Lt Thal, angular gyrus, VLPFC, MTG, and STG |
| | | | | | PPI from rt IFG (decision - draw) [h] | PCC, precuneus |
| | | | | | | Bil visual cortex and VLPFC |

| | | |
|---|---|---|
| | PPI from PCC (draw - decision) h | Lt FEF, Thal, parietal cortex, and angular gyrus |
| | | Rt IPG and MPFC |
| | | ACC and PCC |
| | | Bil visual cortex |
| | | Lt DLPFC |
| | | Rt insula, PMC, paracentral lobule, and Thal |

a Fixed-effect model, voxelwise family-wise error (FWE) rate $p<0.05$

b Uncorrected $p<0.001$

c Uncorrected $p<0.001$ or cluster FWE $p<0.05$

d Voxelwise FWE $p<0.05$

e Constraint principal component analysis, repeated measures analysis of variance (ANOVA)

f ANOVA on multivariate statistics

g Voxelwise FWE $p<0.05$ (Bonferroni)

h Voxelwise false discovery rate $q<0.05$

**Abbreviations**

ACC, anterior cingulate cortex; Bil, bilateral; DLPFC, dorsolateral prefrontal cortex; DMN, default mode network; DTD, draws to decision; FEF, frontal eye field; fMRI, functional magnetic resonance imaging; GB, globus pallidus; IFG, inferior frontal gyrus; IPG, inferior parietal gyrus; IPL, inferior parietal lobule; Lt, left; MB, midbrain; MFG, middle frontal gyrus; MPFC, medial prefrontal cortex; MTG, middle temporal gyrus; OFC, orbitofrontal cortex; PCC, posterior cingulate cortex; PFC, prefrontal cortex; PMC, premotor cortex; PPI, psychophysiological interaction; ROI, region of interest; Rt , right; SMA, supplementary motor are; SPL, superior parietal lobule; STG, superior temporal gyrus; Str, striatum; Thal, thalamus; VLPFC, ventrolateral prefrontal cortex.

**Supplementary Table 2. fMRI studies applying the beads task or its modification in patients with schizophrenia/psychosis.**

| Study | Beads task | Response | Reward /loss | Feedback | Contrast | Brain region |
|---|---|---|---|---|---|---|
| Krug et al, 2014 | Classical | DTD | No | No | Interaction between diagnosis x (beads task - control) [a] | Precuneus<br>Bil MFG<br>Rt IPL, supramarginal gyrus, SFG and IFG |
| Rausch et al, 2014 | Modified (Fish task) | DTD | Yes/No | Yes/No | HC > SCZ for (Decision - draw) [b] | VTA<br>Rt VSt |
| Andreou et al, 2018b | Modified (Box task) | DTD | No | No | Interaction between (Pre - post MCT) x (decision - draw) x (bead ratio) | ROI (task positive network) |

a Clusterwise family-wise error (FWE) rate = $p < 0.05$, cluster forming threshold of $p = 0.001$

b Small volume correction for VTA and VSt

**Abbreviations**

Bil, bilateral; DTD, Draws to decision; fMRI, functional magnetic resonance imaging; HC, healthy controls; IPL, inferior parietal lobule; IFG, Inferior frontal gyrus; MCT, metacognitive training; MFG, Medial frontal gyrus; ROI, region of interest; Rt, right; SCZ, schizophrenia; SFG, Superior frontal gyrus; Vst, ventral striatum; VTA, ventral tegmental area

**Supplementary Table 3. Acquisition parameters for each scan.**

| | Resting fMRI | Structural MRI | Diffusion MRI | Field map |
|---|---|---|---|---|
| Eyes | Open | | | |
| Sequence | GRE-EPI | MPRAGE | SE-EPI | GRE |
| GRAPPA | | 2 | | |
| Orientation | Axial | Axial | Axial | Axial |
| PE direction | A-P | R-L | A-P | R-L |
| TR (ms) | 2000 | 2000 | 10500 | 511 |
| TE (ms) | 30 | 4.38 | 96 | 5.19 and 7.65 |
| TI (ms) | | 990 | | |
| Flip angle | 90 | 8 | | 60 |
| FOV (mm) | 192 x 256 | 225 x 240 | 192 x 192 | 192 x 192 |
| Resolution (mm, x/y/z) | 4/4/4 | 0.94/0.94/1 | 2/2/2 | 3/3/3 |
| Slices | 30 | 208 per slab | 70 | 46 |
| Slice order | Interleaved | | Interleaved | Interleaved |
| Slice gap (mm) | 0 | | | |
| volumes | 180 | | 90 | |
| b=0 volumes | | | 9 | |
| b=1500 directions | | | 81 | |

**Abbreviations**

A-P, anterior to posterior; EPI, echo-planer imaging; fMRI, functional magnetic resonance imaging; FOV, field of view; GRAPPA, generalized autocalibrating partially parallel acquisitions; GRE, gradient-echo; MPRAGE, magnetization-prepared rapid gradient echo; PE, phase encoding; R-L, right to left; SE, spin-echo; TE, echo time; TI, inversion time; TR, repetition time

**Supplementary Figure 1. Trend-level correlations between gray matter/white matter ICs and medication.**

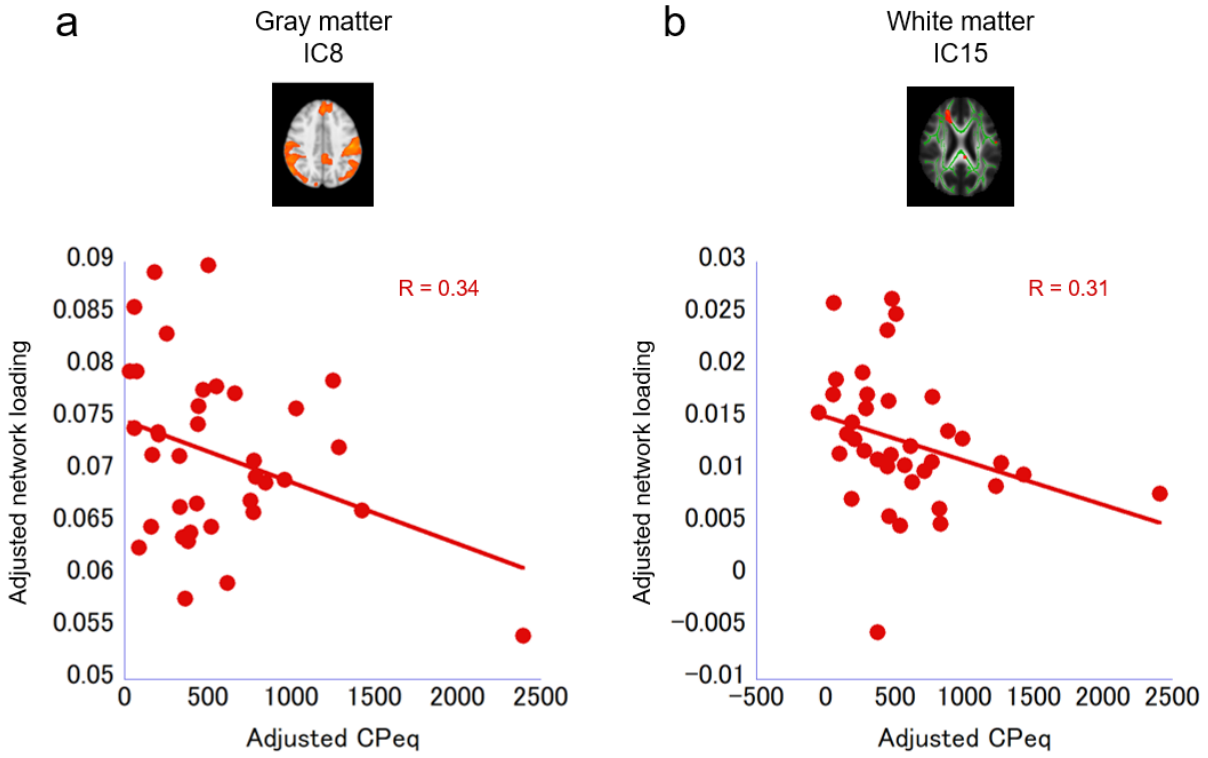


p=0.051 and 0.08 for gray matter IC8 and white matter IC15, respectively (corrected for contrasts). The loading values and CPeq were adjusted for age, sex, JART, and tSNR (for white matter).

**Abbreviations**

CPeq, chlorpromazine equivalent; IC, independent component; JART, Japanese Adult Reading Test; tSNR, temporal signal-to-noise ratio.
